# Network subtypes of cortical similarity reveal molecular correlates of normative and compensatory ageing associated with longevity-gene expression

**DOI:** 10.1101/2025.10.19.25337876

**Authors:** Venia Batziou, Alexandra Young, Timothy Rittman, Vesna Vuksanovic

## Abstract

**Background:** Ageing is marked by widespread cortical changes, but the molecular underpinning and network-level mechanisms associated with this variability remain poorly understood.

**Methods:** We analysed structural MRI data from 952 adults aged 18–94 using morphometric similarity networks, subtype and stage inference, and cortical transcriptomics. Intranetwork connectivity was examined within the three cortical networks and compared with cortical expression patterns of longevity-associated genes.

**Results:** Based on distinct intra-network morphometric similarity patterns and their associations with longevity genes, two ageing-related subtypes emerged. The *normative-ageing* subtype (metabolic-immune) showed connectivity profiles consistent with typical age-related decline and was enriched for genes involved in metabolism, insulin signalling, and immune regulation. The *compensatory* subtype (stress-repair) displayed more preserved intra-network connectivity and was associated with genes linked to stress response, DNA repair, and proteostasis. Although the two subtypes overlapped in oxidative stress and neurodegeneration pathways, they showed divergent molecular signatures associated with different cortical ageing patterns.

**Conclusion:** By integrating network-based morphometry with transcriptomics, we establish a modelling approach that distinguishes normative decline from compensatory adaptation in cortical ageing and provides biologically informed markers of brain ageing.

**Plain Language Summary:** People’s brains do not age in the same way, yet the biological reasons for these differences remain poorly understood. By combining measures of brain-network organisation – estimated using brain MRIs – with maps of gene expression across the cortex, we identified two distinct patterns of healthy brain ageing. One pattern was associated with typical age-related changes in brain networks, while the other reflected altered network organisation and molecular pathways linked to brain cell stress and repair. The two patterns were also associated with different groups of genes involved in metabolism, immune function, stress response, and cellular health. To our knowledge, this is among the first studies to link large-scale brain-networks with ageing-associated genes, highlighting different pathways associated with individual differences in brain ageing patterns.

## Introduction

Ageing is associated with widespread changes in brain structure and function, which unfold along similar, yet heterogeneous trajectories across individuals. Understanding the biological underpinnings of this variability is critical to differentiating healthy ageing from age-related neurodegenerative processes. While neuroimaging studies have delineated normative patterns of cortical morphometry, including age-related changes in cortical thickness, surface area and folding (1–3), how these changes are organised at the systems level and relate to molecular ageing mechanisms remains unclear. Morphometric similarity networks (MSNs) offer a biologically relevant, data-driven framework to investigate coordinated anatomical organisation across the cortex. MSNs take into account the pairwise similarity of multiple morphometric features between cortical regions, providing a meso-scale map of structural connectivity that reflects underlying cytoarchitecture and shared developmental or genetic influences (4– 6). Morphometric similarity is increasingly recognised as a biologically meaningful marker of individual differences in brain anatomy in both health (5, 7) and disease (8–10). The strong association between morphometric similarity and cortical cytoarchitecture suggests that MSNs capture coordinated cellular architecture (11, 12), reinforcing their relevance for ageing research. Furthermore, regions with higher morphometric similarity are more likely to be structurally and functionally connected (13, 14), making MSNs particularly appropriate for studying large-scale brain organisation. In this context, we focused on the default mode network (DMN) and central executive network (CEN), two cognitive networks that support higher-order cognitive processes and are especially susceptible to age-related reorganisation and cognitive decline (7, 15, 16). In addition, we included the visual network as a primary sensory network to provide a contrasting pattern of age-related change and to assess whether subtype-specific effects were preferentially expressed in association rather than sensory systems (7, 15, 16).

Recent evidence suggests that variability in MSNs is, at least in part, shaped by genetic factors (12, 17), supporting their application in genetically informed models of interindividual differences in brain ageing. Moreover, ageing and age-related cognitive decline are highly heterogeneous processes, influenced by metabolic, immune, stress-response, and repair mechanisms acting across the lifespan (18–20). Given their biological and functional relevance, MSNs provide a promising model for characterising this heterogeneity and mapping distinct patterns of cortical organisation to underlying molecular and network-level pathways. We hypothesised that individual variability in morphometric similarity profiles across the cortex could reveal biologically distinct cortical ageing patterns and provide insight into the network-level mechanisms underlying healthy and pathological cognitive ageing.

To address this, we utilised structural MRI, cortical transcriptomics, and normative modelling [using the Subtype and Stage Inference (SuStaIn) (21) approach], to study network-based ageing phenotypes based on MSNs in over 900 healthy adults spanning the adult lifespan. Specifically, we applied data-driven subtyping and staging to intra-network MSN connectivity within three cortical networks: the DMN, CEN, and visual network, and we examined their spatial similarity with the cortical expression patterns of longevity-associated genes.

In this study, we identify two biologically distinct cortical ageing patterns characterised by intra-network organisation and associated with distinct molecular patterns involving metabolic processes, immune regulation, stress response, and cellular repair. These findings suggest that human brain ageing follows heterogeneous biological pathways associated with both normative decline and compensatory adaptation.

## Methods

### MRI datasets

MRI data included 952 healthy participants obtained from three MRI data-sharing platforms: The Nathan Kline Institute (NKI) (22), OASIS-1/2 (23, 24) and IEEEOpenhb (25). In addition, a cohort of 38 cognitively impaired (CDR-defined) OASIS-1/2 participants underwent identical preprocessing and network-construction procedures and was used for comparative analyses with the identified ageing sub-types. All data were previously collected, and extensive documentation on the scanner types and protocols used for these studies can be found in (22–25). In summary, 3D-T1-weighted images from each dataset (see details in Supplementary Information section MRI Datasets) were preprocessed on the Swansea University High Performance Computing facility, supported by Supercomputing Wales, using FreeSurfer version 6.0. This employed the *recon-all* pipeline with default parameters for automated cortical surface reconstruction and morphometric feature extraction. All MRI datasets used in this study were obtained from publicly available, de-identified repositories. Ethical approval and written informed consent were obtained by the original study investigators in accordance with the Declaration of Helsinki, as described in the corresponding dataset publications. Please see also Supplementary Information.

### Morphometric Similarity Networks Construction

For each scan, the cortex was divided into 148 regions of the Destrieux Brain Atlas (DBA) implemented in FreeSurfer. The DBA atlas was applied by automatically classifying each vertex on the surface mesh as sulcal or gyral, which were then assigned to 148 regional labels, i.e., 74 in each hemisphere. Eight morphometric features of interest were used: surface area, gray volume, thickness average, thickness standard deviation, mean curvature, Gaussian curvature, folding index and curvature index. Further details of the procedure are provided in Supplementary Information and (7).

Morphometric similarity networks were created by calculating the correlation coefficient of z-normalised morphometric features to create a 148 *×* 148 matrix for each participant. We then applied Fisher’s transformation to convert the correlation coefficients to normally distributed z-scores. Self-correlations were replaced with the value 0. Three networks were created with combinations of different features for each participant: 1) a 4-volumetric-feature (4vf), 2) a 4-curvature-feature (4cf), and 3) a 5-feature (5f) model (7). In short, the 4vf network was constructed on the inter-regional pair-wise correlations between the volume, surface area, thickness, and thickness standard deviation. The 4cf networks were constructed by correlating intra-regional curvatures and their indices; 5-feature networks were constructed on the intra-regional correlations between the volume, surface area, thickness, Gaussian curvature, and folding index. These feature combinations were chosen to capture complementary aspects of cortical morphology that differ in their sensitivity to ageing and developmental influences. The 4vf network combined volumetric and area measures (volume, surface area, thickness, and thickness variability), which are known to decline with age and strongly reflect neurodegenerative and plastic processes (26). In contrast, the 4cf network, based on curvature-derived measures, represents geometric properties of cortical folding that are largely established early in development and relatively stable across adulthood (3). The 5f network integrated both volumetric and curvature-based metrics to provide a more comprehensive morphometric profile (5); however, this combination may dilute age-related variance by combining features with different biological timescales. Thus, examining all three configurations allowed differentiation between structural features that are age-sensitive (4vf), developmentally constrained (4cf), and combined but less specific (5f), providing a more refined assessment of which morphometric combination/dimension best captures ageing-related network (re)organisation.

We calculated weighted node degree (node strength) (27) to quantify within-network connectivity in three cortical networks: two higher-order association networks – the DMN and CEN – and the visual network, a primary sensory network. The mean value of the weighted node degree (node weight) for each network’s left and right hemisphere was used as a proxy for *‘intra-network connectivity’*. These networks are known to show distinct structural changes with ageing, with association networks such as the DMN and CEN exhibiting earlier and more pronounced morphometric changes than primary sensory systems such as the visual network (28, 29). The DMN is associated with self-referential and memory-related processes; the CEN is involved in attention, working memory, and goal-directed control; and the visual network supports sensory-perceptual integration. Changes in within-network and between-network connectivity have been well documented in ageing and neurodegeneration. For example, previous studies have shown that the two main association networks – the DMN and CEN – undergo altered within-network organisation with ageing, while also exhibiting changes in interactions with other cortical systems (28, 30, 31).

### SuStaIn Model

For SuStaIn modelling (21), participants were divided into a young-adult (YA; 18–30 years, n = 199) reference group and a healthy-ageing (HA; 45–94 years, n = 753) group (see Table 1 and Fig.S1 for their demographic characteristics). SuS-taIn is an unsupervised machine learning model that identifies groups of individuals with distinct patterns of biomarker progression and assigns each individual both a subtype and a stage within that subtype (21). This modelling approach was initially applied in the context of neurodegeneration in dementia to ‘stage’ (heterogeneity in) disease progression relative to a healthy (reference) group. That is, the model uses cross-sectional data to infer the most likely ordering of biomarker events (transitions between z-score thresholds) relative to the reference group, with each stage representing the cumulative occurrence of one or more such events rather than a specific period of time. By analogy, here we applied SuStaIn to investigate heterogeneity in healthy brain network-organisation patterns across the adult lifespan, with the aim of identifying distinct patterns and their associated molecular signatures.

**Table 1.** Demographic characteristics of the cohorts.

|  | YA | HA | CI |
| --- | --- | --- | --- |
| Male/Female | 97/102 | 320/433 | 14/24 |
| Age range (years) | 18 -30 | 45 - 94 | 64 - 96 |
| Average age (years) | 22(3) | 66(11) | 78(8) |

Abbreviations: YA - Young Adults; HA - Healthy Ageing; CI - Cognitive Impairment.

In our analysis, the input biomarkers were intra-network MSN-based node-strength values derived for each participant’s network and hemisphere, and normalised relative to the YA reference group. The young-adult group was used only to define a reference pattern of network organisation, as required by the model. In our models, higher stage numbers therefore indicate a later position within the inferred ageing progression pattern, but do not imply a faster rate of ageing or a fixed chronological duration. As SuStaIn models were fitted independently for each network and hemisphere, the number of inferred stages is model-specific. That is, the number of SuStaIn stages reflects the number and ordering of biomarker events supported by each independently fitted model. Stage numbers are therefore interpretable only within their respective models and should not be compared directly across different network or hemisphere models.

Subtype classification and stage inference were performed within the HA group (45–94 years) only. Participants aged 30–45 years (early adulthood) were excluded to create a clearer separation between the reference (young-adult) and healthy-ageing groups and to model ageing-related changes, which were the focus of this study. This choice was informed by previous lifespan neuroimaging studies showing that many cortical measures, including thickness, surface area, and volume, exhibit relatively stable patterns during early adulthood followed by more consistent age-related decline from midlife onwards, with regional variability in the timing and rate of change (32). We included participants from 45 years of age as we wanted to investigate early signs of differentiation in network-organisation patterns. The SuStaIn model was fitted using z-score thresholds of 0.5, 1.0, and 1.5 (maximum z = 3), representing increasing deviations of the biomarkers from the reference distribution. Model selection was based on k-fold cross-validation, in which the data were repeatedly partitioned into training and testing subsets to evaluate model stability and reproducibility across increasing numbers of subtypes. Given the balanced sex distribution across groups, sex was not included in the SuStaIn modelling but was considered in subsequent statistical analyses where appropriate (see Statistical Analysis).

### Genetic Analysis

Ageing-associated genes were extracted from the GenAge human ageing genes database, derived from the latest stable build of GenAge (33). The database, which consists of 307 age-related genes in total, was matched to genes represented in the Allen Human Brain Atlas (AHBA) database, similar to our previous study (34) (see also Supplementary Information for more detailed explanation). This resulted in 251 ageing-related genes common to both databases and the matched dataset used in our longevity-gene-MSNs associations analysis. The full list of genes and their function, and related data files are available in the project’s OSF repository folder. DBA regional gene-expression maps were extracted from AHBA micro-array data. Where a DBA region was not mapped to the AHBA gene expression array, missing values were replaced with the corresponding overall mean, either the whole brain or nodal mean depending also on the statistical test performed on the maps.

To biologically characterise the identified subtypes, partial correlations were computed between regional gene-expression values and subtype-specific node-strength measures across all 148 cortical regions of the DBA, controlling for regional cortical surface area and thickness. Because both morphometric measures are strongly influenced by ageing and vary systematically across the cortex, they were included as covariates to reduce the influence of global anatomical variation on gene–network associations. This approach, adapted from our previous work (35), aimed to identify associations reflecting coordinated network-level organisation rather than broad regional morphometric effects. Multiple comparisons were controlled using false discovery rate (FDR) correction at q < 0.05. In addition to the subtype-specific analyses, gene–MSN associations were also evaluated across all participants pooled together using the same partial-correlation approach, allowing comparison between global and subtype-specific molecular patterns (see Supplementary Information for details).

The functional annotation of significant genes was obtained from RefSeq summaries available through the National Center for Biotechnology Information (NCBI) Gene database, with the resulting annotation table and supporting files available in the project’s OSF repository folder.

### Statistical Analysis

Study participants were divided into young adults (YA) and healthy ageing (HA) groups, according to their age band. Demographic characteristics of the participants for the YA and HA groups are shown in Table 1.

Cortical morphometric characteristics across the study groups were compared using the non-parametric Kruskal-Wallis test followed by the Mann-Whitney U test with Bonferroni correction for multiple comparisons. Where there were no mean differences between two classes, logistic regression was used to test for the effect of age and gender on subtype classification. Associations between network patterns and variation in the expression of longevity genes were estimated using partial correlation, with cortical thickness and surface area as covariates. P values were FDR-corrected at a significance threshold of 0.05.

The overall data analysis pipeline is summarized in Fig. 1.

**Fig. 1.**
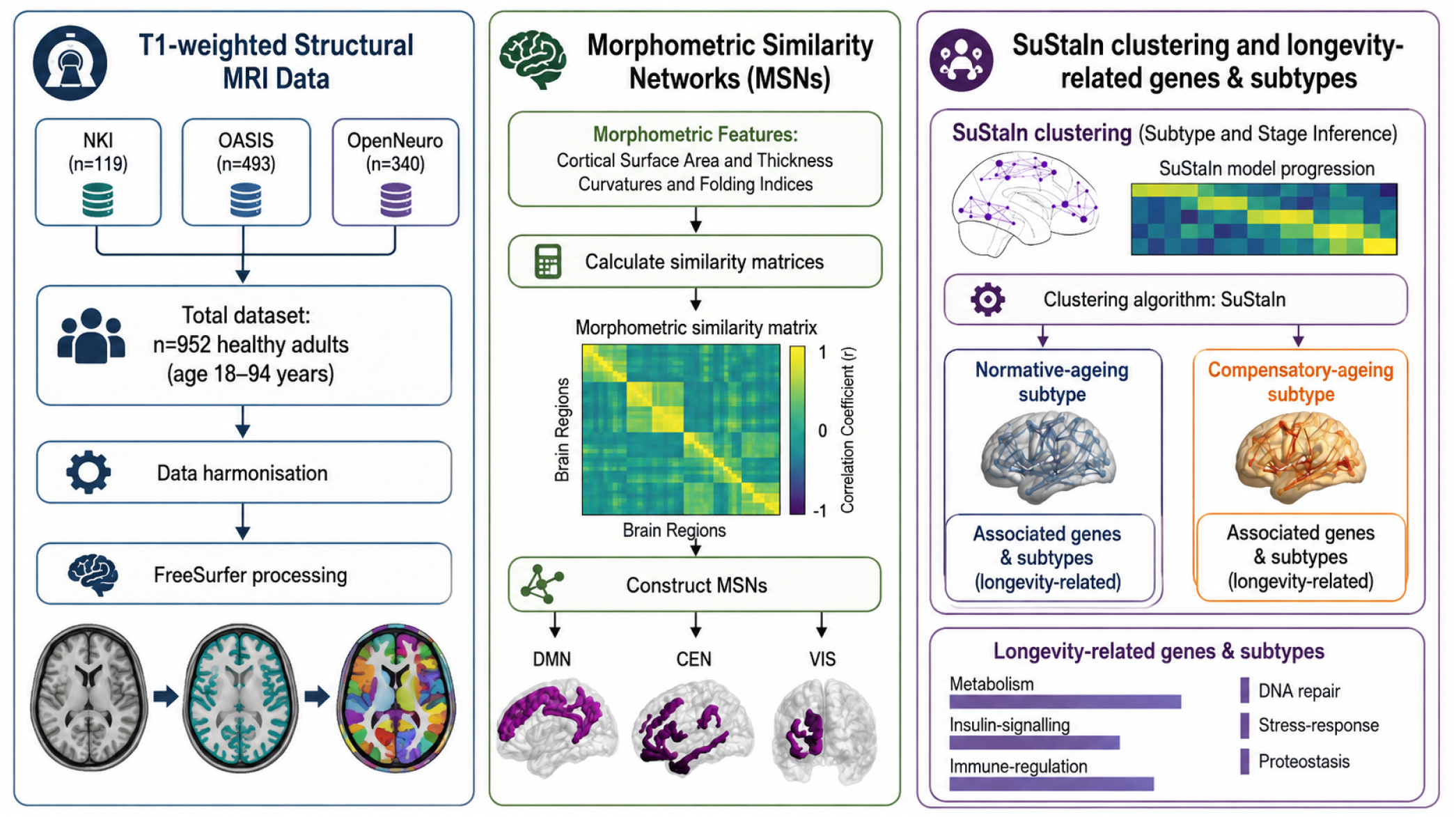
Data analysis pipeline.

## Results

### Age-Associated Subtypes in Morphometric Similarity and Gene Expression

To investigate whether age-related differences in cortical network organisation are reflected in patterns of intra-network connectivity, we applied SuStaIn to intra-network node strength measures derived from the DMN, CEN, and visual networks. Analyses were performed using three MSN configurations: a 5-feature model, a 4-volumetric-feature model, and a 4-curvature-feature model. Given established hemispheric asymmetries in cortical architecture, gene expression, and ageing heterogeneity (36–38), the SuStaIn model was fitted separately for left and right hemispheres. This approach enabled us to assess whether patterns of morphometric similarity and subtype differed between hemispheres, providing a more anatomically and biologically informed characterisation of cortical ageing. We identified two distinct subtypes: Subtype 1, and Subtype 2. These were evident only within the 4vf MSNs, meaning that only the volumetric-feature MSN configuration (4vf) generated stable and separable inferred progression trajectories within the model. In addition, the subtypes exhibited hemispheric asymmetry, i.e., a single subtype was found in the right hemisphere for the visual network. This is consistent with the view that visual cortices are the last to experience age-related changes compared to, for example, frontal or temporal regions (16, 39). Subsequent gene-expression analyses revealed distinct molecular associations for each sub-type, enabling more intuitive, biological, interpretation as the *normative-ageing* (Subtype 1) and *compensatory* (Subtype 2) subtypes. Fig. 2 visualises the number of individuals within each subtype and overlaps across the DMN, CEN and visual networks. Subtype assignments were highly consistent across networks, with more than 80% of individuals retaining the same subtype membership regardless of the network examined.

**Fig. 2.**
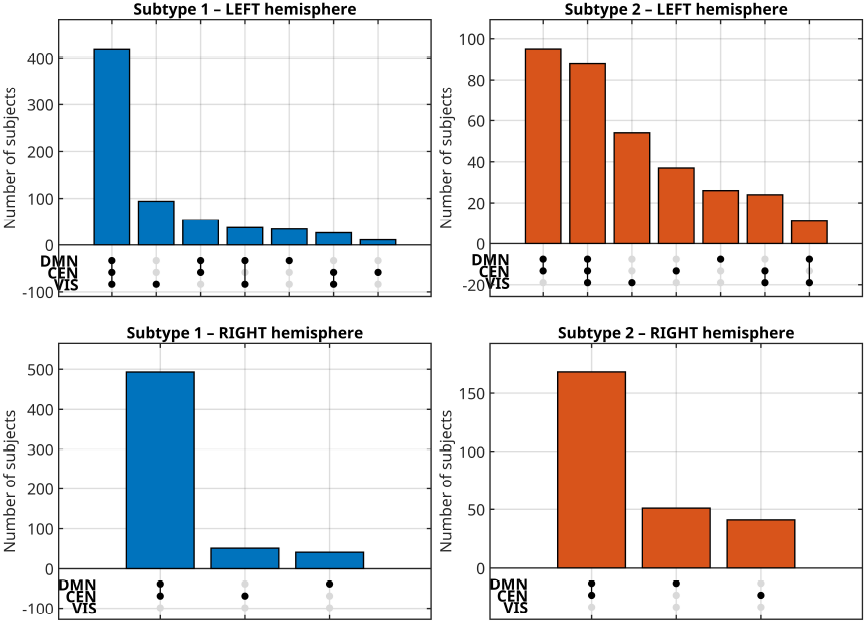
Visualisation of subtype overlap across the Default Mode (DMN), Central Executive (CEN), and Visual (VIS) networks in the left and right hemispheres. Bar heights represent the number of participants exhibiting each combination of subtype membership across the three networks. Filled dots identify the networks contributing to a given overlap pattern.

The mean age across SuStaIn *stages* for the subtypes, within the DMN, CEN and visual (left to right panels) networks is shown in Fig. 3. Subtype 2 consistently shows higher mean ages than Subtype 1, across DMN and CEN intranetwork connectivity, while in the visual network, age differences were less pronounced. Across networks, higher SuStaIn stages corresponded to older mean ages, indicating age-related progression in intra-network connectivity (see also Fig.S7). Visualisations of the subtypes’ associations with age are shown in Fig.S4. Both subtypes show that morphometric similarity increases with age across the DMN, CEN, and visual networks. This pattern indicates that cortical regions within each network become more morphometrically ‘similar’ to one another, reflecting coordinated structural (morphometric) change with ageing. In addition, *compensatory* subtype (Subtype 2), had a higher mean age compared to *normative-ageing* subtype (Subtype 1) (70.0 vs 65.5 years). However, the group means did not differ significantly. In the repeated test where age was included as a covariate, the results showed significant (p<.0001) contribution of age to the subtype classification, reaching the accuracy of ≈93% (see Fig.S3).

**Fig. 3.**
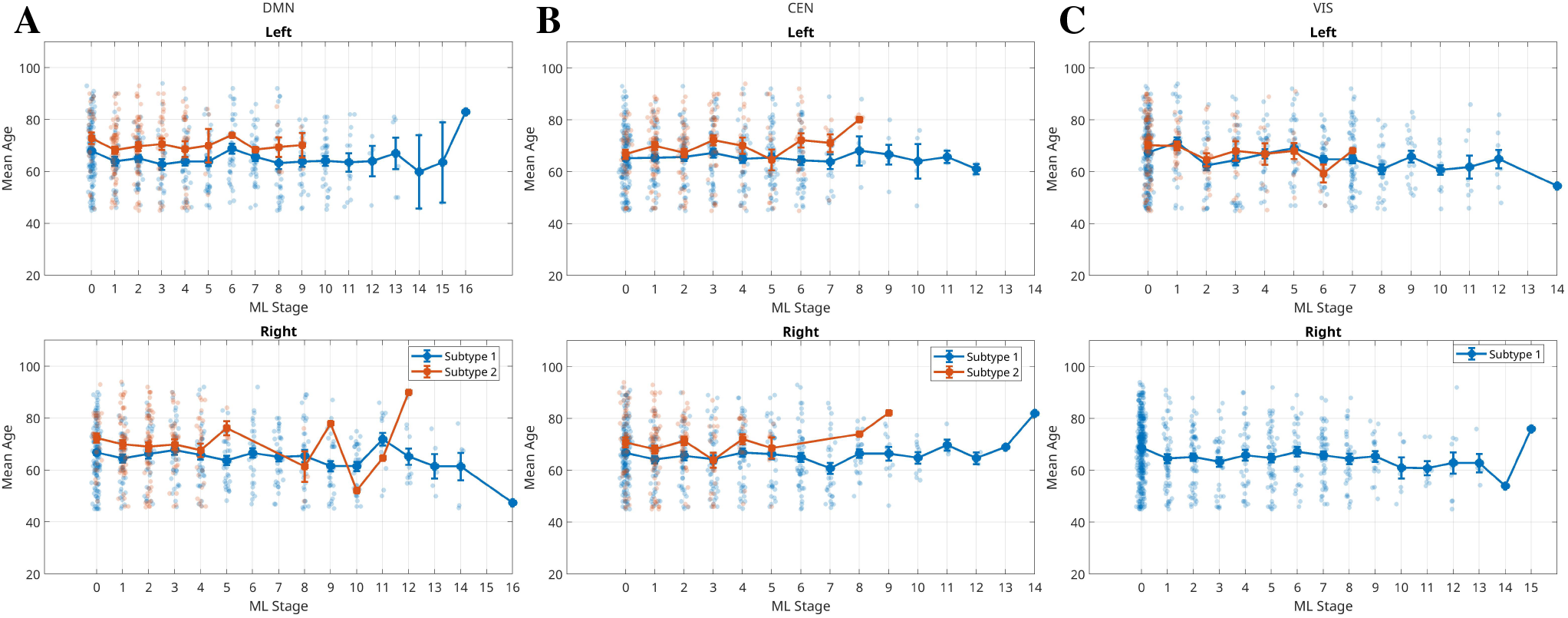
Mean age across SuStaIn stages. Subtype 1 (*normative-ageing*) and Subtype 2 (*compensatory* ) mean age within the DMN, CEN, and visual networks (left to right panels). Across networks, participants assigned to Subtype 2 consistently showed higher mean ages than those assigned to Subtype 1. The separation between subtypes was most pronounced in the DMN and CEN, whereas age differences between subtypes were less evident in the visual network. DMN - Default Mode Network; CEN - Central Executive Network; VIS - Visual Network.

#### Associations between Network Subtypes and Longevity Genes

Having established two intra-network connectivity subtypes, we next investigated whether these distinct patterns of connectivity are associated with cortical maps of gene expression. In particular, we were interested in any significant associations between intra-network connectivity and human longevity genes (AgeGene) (33).

To examine the relationship between morphometric similarity and genes implicated in human longevity (33) ageing-gene–MSN associations were calculated using partial correlation, while controlling for regional variation in surface area and cortical thickness. Although a similar number of longevity-associated genes were significantly correlated with morphometric similarity across subtypes (Fig. 4), their spatial and network-level profiles differed. In Subtype 1, morphometric similarity was associated with the expression of genes involved in, or related to metabolism, insulin signalling, and immune regulation. These included *IRS1* and *PPARGC1A* (glucose/insulin pathways and cholesterol/obesity), *GSK3B* (energy metabolism and neurodegeneration), and transcriptional regulators (*E2F1, MXD1*). Notably, *FOXO1* (myogenic growth, differentiation) and *PLCG2* (immune signalling, autoinflammation) showed negative associations with intra-network connectivity, suggesting potential protective or resilience-related effects in this subtype. Subtype 2 was characterised by genes involved in cell stress, apoptosis, DNA repair, and protein degradation. These included *DDIT3* (ER-stress–induced apoptosis), UBB (ubiquitin system, Alzheimer’s/Down syndrome), *STUB1* (protein quality control, spinocerebellar ataxia), and GSK3B (metabolic and neurodegeneration pathways). However, genes such as *FEN1*, a key DNA repair gene, showed a negative association, indicating possible protective contributions against progression in this subtype. Despite overlaps (E2F1, *MXD1*, GCLM, *GSK3B*), Subtype 2 displayed stronger links to stress-response and processes closely tied to neurodegeneration [see, e.g., (40, 41)]. Overlapping genes – E2F1, *MXD1*, GCLM, *GSK3B* – mainly clustered around cell cycle regulation, oxidative stress, and metabolic/neurodegenerative pathways. This suggests a common core biology across both subtypes, with divergence in additional stress/apoptosis vs. metabolic/immune signatures. Genes unique to Subtype 1 (*normative-ageing*) were involved in metabolism, insulin signalling, mitochondrial function, and immune regulation, while genes unique to Subtype 2 (*compensatory*) were related to cell stress responses, apoptosis, DNA repair, and protein degradation – processes closely tied to neurodegeneration [see, e.g., (40, 41)].

**Fig. 4.**
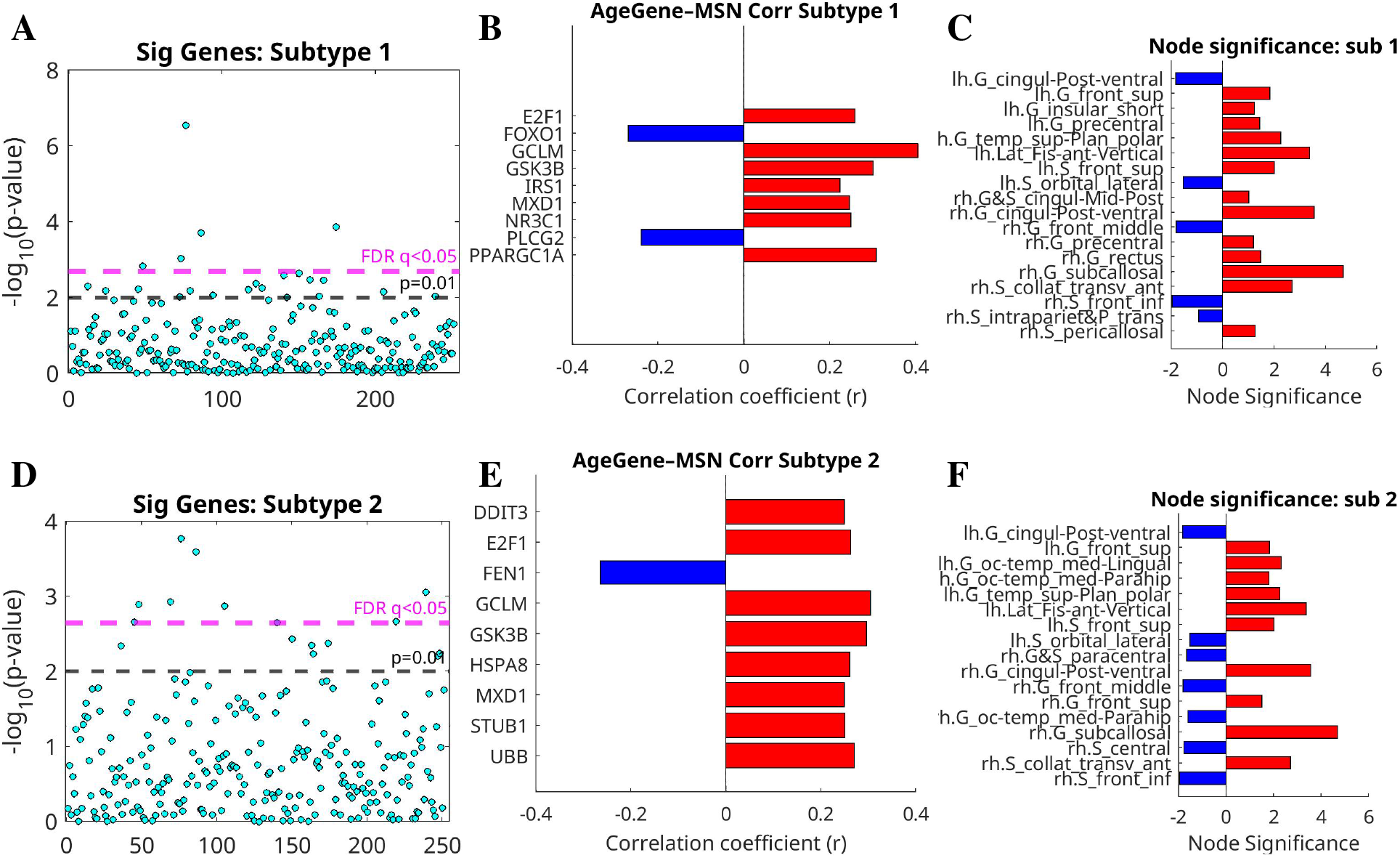
Association between expression of longevity genes and cortical morphometric similarity across ageing subtypes. **Upper Panels (A–C):** Significant gene-MSNS associations for Subtype 1 (*normative ageing subtype*). **Lower Panels (D–F):** Association between longevity-gene expression and cortical morphometric similarity across ageing subtypes. Partial correlations were computed across 148 cortical regions of the DBA while controlling for regional surface area and its thickness. Statistical significance was assessed using two-sided partial correlation tests and corrected for multiple comparisons using the false discovery rate (FDR; q *<* 0.05). Associations significant at the uncorrected threshold (P *<* 0.01) are also shown. Middle Panels (B/E): Significant gene-MSNs associations. Statistical significance is shown at an FDR-corrected threshold of 0.05 and an uncorrected threshold of 0.01, with associations controlled for regional variations in cortical thickness and surface area. Subtype 1 was enriched for genes involved in metabolic regulation and immune processes (e.g., *IRS1, PPARGC1A, GSK3B, E2F1*), with *FOXO1* and *PLCG2* showing negative associations suggestive of protective effects. Subtype 2 was characterised by stress- and neurodegeneration-related genes (e.g., *DDIT3, UBB, STUB1, GSK3B*), with *FEN1* showing a negative association consistent with possible protection. Overlapping genes (*E2F1, MXD1, GCLM, GSK3B*) indicated shared pathways, but Subtype 2 showed stronger links to stress-response mechanisms, contrasting with the more metabolic/immune profile of Subtype 1. Right panels: Cortical regions contributing significantly to the gene–MSN associations *normative-ageing* (Subtype 1) vs *compensatory* (Subtype 2) groups. Bar plots show cortical regions with significant associations between intra-network morphometric similarity and longevity-genes-expression profiles.

To test the robustness of the ageing-gene–MSN associations, we re-ran the analyses without controlling for cortical thickness and surface area. The resulting set of significant genes was different (Fig.S5), suggesting that some associations may be driven primarily by global cortical morphology. Thus, our results corrected for the morphometric confounds (regional cortical thickness and surface area) more accurately reflect how regional morphometric similarity is associated with the expression of longevity genes.

#### Longevity Genes and Subtype-specific Node Maps

To better understand the regional contributions to ageing-gene–MSN associations, we identified the top contributing cortical nodes for each subtype. Fig. 4 (right panels) shows node significance in ageing-gene-MSNs associations, calculated on partial correlations corrected for variations in regional surface area and its thickness. There were 18 nodes for the *normative-ageing* Subtype 1, and 17 nodes for the *compensatory* subtype (Subtype 2). Eleven nodes were shared between the two subtypes, six in the left hemisphere and five in the right. Five shared nodes belonged to the DMN: bilateral posterior ventral cingulate, left superior frontal gyrus, left superior frontal sulcus and right middle frontal gyrus. The right middle frontal gyrus also belonged to the CEN. The concentration of left-sided overlapped regions in the DMN system suggests possible hemispheric differences in the associations between longevity-gene expression and cortical network organisation.

The remaining nodes were subtype-specific. Subtype-specific regions included the left insular gyri, bilateral precentral gyri and right intraparietal sulcus in Subtype 1 *normative-ageing*. Subtype 2-specific regions included the left lingual gyrus, bilateral parahippocampal gyri, right superior frontal gyrus, right paracentral region and right central sulcus, involved in the DMN and in sensorimotor domains. The *compensatory* subtype (Subtype 2) uniquely involved medial occipital and temporal regions including the lingual and parahippocampal gyri, suggesting greater involvement of visual and memory-related circuits. Six Subtype 1 nodes and nine Sub-type 2 nodes belonged to the DMN, CEN or visual network combined, leaving 12 and eight nodes, respectively, outside these network definitions. Network-affiliated nodes were distributed equally between hemispheres in Subtype 1 (three left, three right), while Subtype 2 included five left-sided and four right-sided nodes. Regions involved in the subtypes were balanced between hemispheres: *normative-ageing* sub-type (Subtype 1) 8 left, 10 right; *compensatory* subtype (Sub-type 2) 8 left, 9 right.

#### Genetic Associations with Bilateral Cortical Morphometric Networks

Finally, to explore the broader molecular underpinnings of cortical network organisation, we examined associations between bilateral MSNs and cortical gene-expression maps from the AHBA resampled to the DBA resolution. This analysis included all genes from the AHBA (mapped onto the DBA) to capture global transcriptional influences on cortical morphometric similarity beyond longevity-related pathways. The results revealed pronounced hemispheric asymmetry in gene–MSN associations, explaining 24% of variance in the left hemisphere and 10% in the right (Fig. 5). These associations were consistent across both age groups, suggesting that left-lateralised molecular-structural coupling represents a stable feature of cortical organisation across the adult lifespan.

**Fig. 5.**
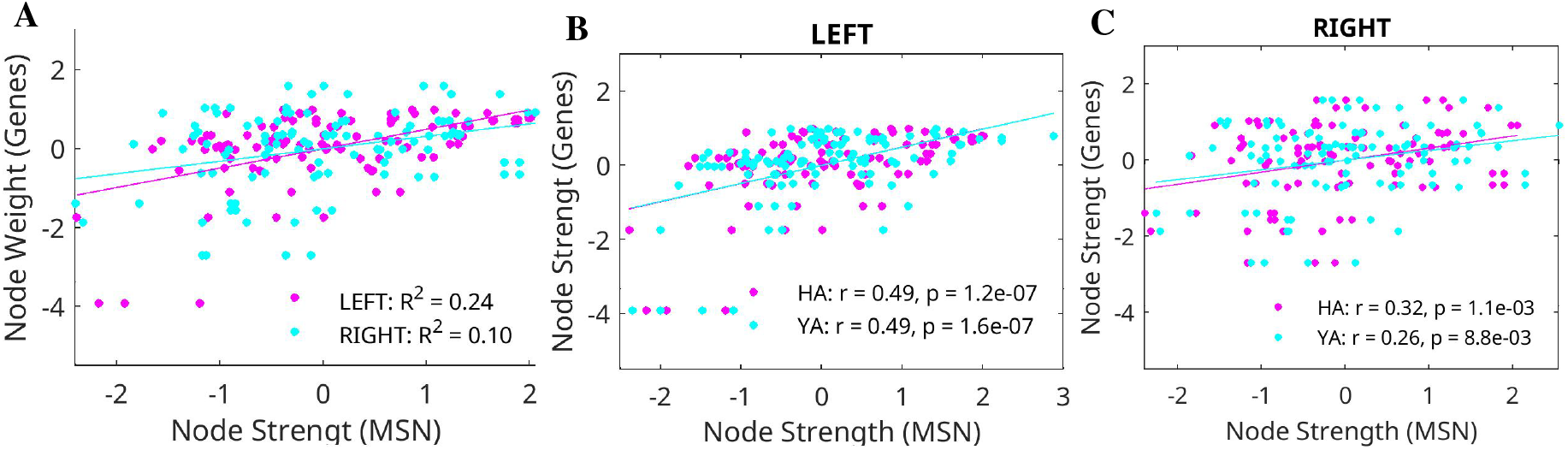
Coupling between cortical network organisation and longevity-gene expression. Scatter plots show the relationship between MSN node strength and regional longevity-gene expression across 148 cortical regions in the left and right hemispheres (74 each), and YA (n=199) and HA (n=753) groups (**A–C**). Solid lines indicate least-squares linear regression fits. Pearson’s correlation coefficient (*r*) and the corresponding two-sided P value are shown in each panel. The proportion of variance explained (R^2^) is also reported.

### Intra-network Connectivity and Ageing

Given that our age-related subtypes of intra-network connectivity were found only for 4vf MSNs, we next investigated whether intranetwork connectivity differed between young adults and the healthy ageing group for 5f and 4cf MSNs. Intra-network node strength was calculated for the DMN, CEN, and visual networks of the three different MSNs (4vf, 4cf and 5f) and statistical differences were analysed between YA and HA groups. Fig.S2 shows significant group differences in all three cognitive networks for the 5f network, and within DMN for the 4cf. This is in agreement with (5), where 5f MSNs were compared at the group-averaged level. Importantly, while the YA group showed clear differences between 5f and 4vf networks, these distinctions were absent in the HA group, suggesting a weakening of network-specific integration with age.

We then assessed broader morphometric changes across the cortex. Fig. 6 shows average morphometric features in YA and HA groups. All features, except the two curvature metrics, were significantly reduced in the HA group. These findings were consistent across bootstrapped analyses.

**Fig. 6.**
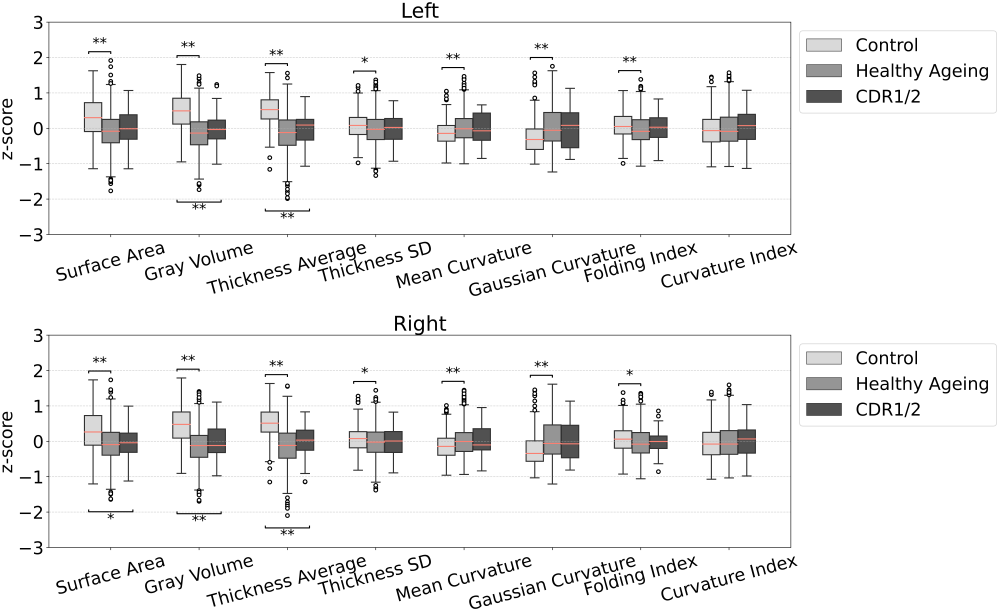
Morphometric changes across cortical features. Boxplots show healthy young adults and healthy ageing group z-normalised values to the corresponding feature mean. Mann-Whitney U test with Bonferroni correction. (**: p-value *<* 0.001 *: p-value *<* 0.01.) L = Left, R = Right hemisphere. YA = Young Adults, HA = Healthy Ageing

### Relationship Between Subtypes and Cognitive Impairment

Finally, to contextualise the ageing-related subtypes, we compared HA subtypes to cognitively impaired individuals classified by the Clinical Dementia Rating scale (CDR=1 or 2). Figure 7 shows that, on average, the *normative-ageing* subtype (Subtype 1) shows a pattern of intra-network organisation more similar to that observed in the CDR1/2 group, suggesting that both groups share comparable network-level structural changes typical of age-related decline. In contrast, the *compensatory* subtype (Subtype 2) shows a distinct profile, characterised by intra-network similarity patterns that differ from both the *normative-ageing* and YA groups. This suggests that *compensatory* MSN reorganisation may represent a distinct form of cortical ageing rather than simply more advanced normative ageing.

**Fig. 7.**
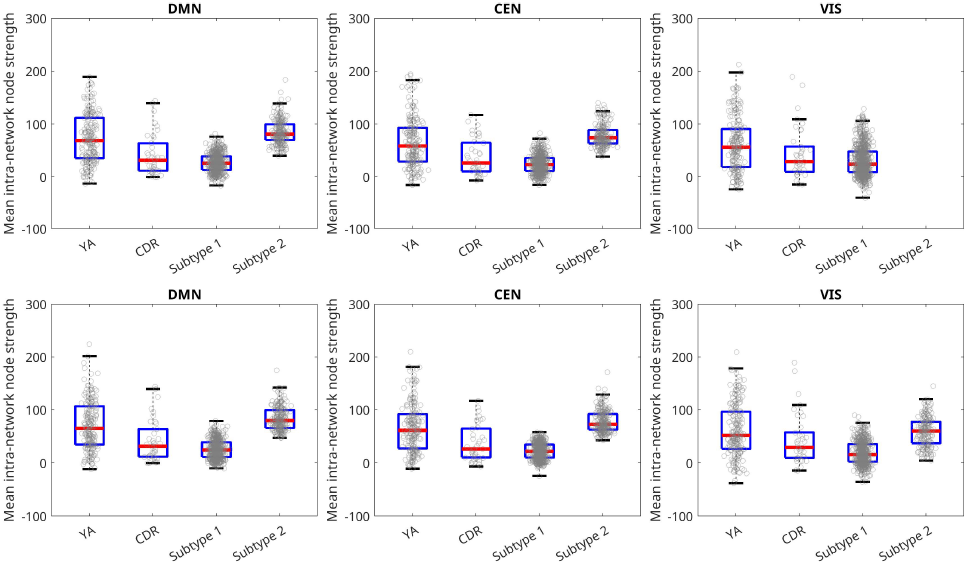
Mean intra-network node strength for the default mode (DMN), central executive (CEN), and visual (VIS) networks in young adults (YA; n =199), healthy-ageing Subtype 1 (*normative-ageing*) and Subtype 2 (*compensatory* ) groups, and individuals with mild-to-moderate cognitive impairment (CDR1/2; n=38). **Upper panels**: results for the right hemisphere. **Lower panels**: results for the left hemisphere. Group differences were assessed using the two-sided Mann–Whitney U test with Bonferroni correction for multiple comparisons. Exact P values are provided in the figure. Significant differences were observed between Subtype 2 and the CDR1/2 group for all networks in which Subtype 2 was identified. No participants were assigned to Subtype 2 in the right-hemisphere VIS network; therefore, no corresponding boxplot is shown. Participant count for each network and hemisphere is shown in Fig. 2.

The CDR1/2 group’s broader morphometric changes across the cortex are shown along the YA and HA groups (see Fig. 6).

## Discussion

Here we present, to our knowledge, the first investigation of associations between longevity-related gene expression and morphometric similarity networks in a large adult cohort of over 900 individuals spanning the adult lifespan. The study design, integrating morphometric similarity network analysis with cortical maps of longevity-gene expression and normative modelling (incorporating subtyping and staging), provides a robust approach for capturing individual variation in brain network organisation across adulthood. This approach revealed two distinct morphometric similarity subtypes, characterised by unique as well as overlapping associations with cortical expression patterns of ageing-related genes. In addition, we linked these associations to three major cognitive networks; the default mode, central executive, and visual networks. Based on their combined network and molecular characteristics, we refer to these as the normative-ageing and compensatory subtypes, reflecting distinct but complementary patterns of cortical ageing. The normative-ageing subtype was predominantly associated with metabolic and immune-related pathways, whereas the compensatory sub-type was characterised by genes involved in stress response, cellular repair, and proteostasis. In addition, we found evidence of hemispheric asymmetry, where gene-MSN coupling accounted for approximately 24% of the variance in the left compared with 10% in the right hemisphere.

These two distinct age-related subtypes of intra-network morphometric similarity arose from volumetric-feature-based MSNs (4vf) and were present across the three cognitive networks: DMN, CEN, and visual (Figs. 2 & 3). The sub-types exhibited robust consistency across individuals and networks, as well as hemispheric asymmetries, particularly in the visual system. The presence of a single visual subtype in the right hemisphere aligns with prior evidence suggesting later vulnerability of the visual cortex to ageing compared to other higher-order association areas of the cortex. The lack of such subtype separation from the 5 feature (5f) MSNs or curvature-only MSNs (4cf), indicates agreement of our results with the current view of the field. That is, that volumetric morphometric features are more sensitive to age-related network changes (29). The 5f MSNs, while a comprehensive combination of features, may dilute ageing-relevant signals by combining age-sensitive volumetric measures with relatively age-invariant curvature features. In contrast, the 4cf networks based solely on cortical folding, likely reflect more developmentally fixed aspects of brain structure that are less responsive to changes with ageing [for a review see (42)]. This highlights the specificity and sensitivity of volumetric metrics (regional surface area, thickness and volumes) in capturing biologically meaningful heterogeneity in cortical ageing.

Among our main findings is the differential spatial association of the two subtypes with cortical maps of longevity-gene expression (Fig. 4B, E). By incorporating regional gene-expression information to annotate differences in MSN organisation, these findings add a molecular dimension to the network heterogeneity identified by SuStaIn and may provide insight into the biological processes underlying these variations. The *normative-ageing* subtype (Subtype 1) was spatially associated with genes involved in circadian regulation, glucocorticoid signalling and metabolic processes, pointing to pathways broadly implicated in neuroendocrine ageing and resilience (43). This interpretation is consistent with large-scale, pathway-wise genomic studies of brain and systemic ageing that report associations with lipid-metabolic and immune processes, together with drug-target associations, broadly aligning with the metabolic–insulin and immune profile of this subtype (44, 45). In contrast, the *compensatory* subtype (Subtype 2) was associated with genes involved in DNA repair and proteostasis, suggesting a molecular profile related to the maintenance of genomic and proteomic integrity. These profiles were not mutually exclusive: four genes – *GCLM, E2F1, MXD1* and *GSK3B* – were shared across the two subtypes, indicating common processes involving oxidative stress and apoptosis that may contribute to general brain ageing. Thus, while the subtypes share some ageing-related molecular processes, they also show distinct molecular associations that may underlie differences in their structural network patterns. When considered alongside their network characteristics (Fig. 7), these molecular findings support their interpretation as *normative-ageing* and *compensatory* subtypes. In particular, Subtype 1 showed greater similarity to the cognitively impaired group, whereas Sub-type 2 showed higher intra-network connectivity than both the young and CDR1/2 groups.

The regional distribution of the gene–MSN associations provides an anatomical context for the shared and overlapping molecular profiles described above (Fig. 4C, F). Regional differences in these associations may help explain the biological heterogeneity—here, variation in longevity-gene expression across the brain—associated with the intra-network connectivity patterns identified by SuStaIn. At the same time, shared involvement of key DMN hubs in the posterior cingulate and frontal regions, together with the CEN-associated middle frontal hub, suggests that the two subtypes retain a common core network organisation. This supports their interpretation as variants of the same broader pattern rather than entirely separate patterns. Consistent with the modular yet integrated organisation of the cerebral cortex, these hubs support interactions between spatially distributed and functionally specialised cortical regions. DMN hubs contribute to internally directed processes, including self-referential thought, memory and future-oriented cognition (46, 47), whereas the CEN-associated prefrontal hub helps coordinate information across cortical systems and flexibly adjusts their interactions according to current goals and task demands (48). Their shared involvement therefore provides a common anatomical basis, while the wider regional distribution and molecular profiles distinguish the two subtypes. In the *normative-ageing* sub-type, involvement of the left short insular gyri suggests an association with salience processing (49). Sensorimotor involvement was evident in both subtypes but differed in its regional distribution. The *normative-ageing* subtype included the bilateral precentral gyri, with additional right intraparietal involvement suggesting a broader visuomotor contribution (50, 51), whereas the *compensatory* subtype included the right paracentral and central-sulcus regions, supporting medial and peri-central sensorimotor involvement (52, 53). Shared cingulate and subcallosal involvement further indicates common limbic components. Together, these findings suggest that the molecular associations of both subtypes extend across systems supporting cognitive function and coordinated responses. The additional visual and medial temporal involvement in the *compensatory* subtype, including the bilateral parahippocampal gyri, a DMN-associated hub (54), suggests links with perceptual and memory-related systems (47). This regional pattern complements the stress-response and repair-related molecular profile, although the involvement of these regions does not, by itself, show that their functions are preserved or that the observed changes are compensatory. Associations outside the DMN, CEN and visual network nodes further indicate that the molecular correlates of these ageing patterns extend beyond the selected network boundaries. This interpretation is consistent with structural covariance and morphometric network studies showing that age-related cortical differences involve anatomically distributed systems (35, 55, 56).

The observed increases in intra-network morphometric similarity may reflect convergent structural changes across regions within the same functional networks. Such convergence could represent shared susceptibility to ageing processes or coordinated adaptation (35, 55, 56). The contrasting molecular associations provide context for interpreting these patterns as normative-ageing and potentially compensatory forms of cortical reorganisation, with the latter associated with genes involved in stress responses and cellular repair. Separately, our hemisphere-level analysis showed stronger gene–MSN coupling in the left hemisphere, explaining approximately 24% of variance compared with 10% in the right (Fig. 5). This pattern suggests possible hemispheric differences in the spatial correspondence between gene expression and cortical network organisation, which may relate to established links between functional lateralisation and structural connectivity (37).

Finally, we contextualised these subtypes by comparing them to individuals with mild to moderate cognitive impairment (CDR1/2). The two cortical ageing subtypes revealed by our analyses capture distinct yet complementary trajectories of brain ageing. The normative-ageing subtype, although on average younger than the compensatory group, exhibited morphometric network profiles resembling those observed in individuals with early impairment. This suggests a trajectory characterised by more homogeneous structural change in typical (normative) age-related decline. This normative pattern was associated with the expression of longevity-related genes involved in metabolism, immune regulation, and neuroendocrine signalling (57–59). In contrast, the compensatory subtype showed greater intra-network similarity across association cortex and was enriched for genes supporting DNA repair and proteostasis, consistent with an adaptive, stress-response–driven pathway (60–62). Because increased morphometric similarity can arise from both coordinated tissue loss and adaptive network reorganisation (35, 55, 56), the biological interpretation of this pattern is informed by the associated gene-expression profiles, which were enriched for stress-response, DNA-repair, and proteostatic pathways (Fig. 4). Together, these findings indicate that age alone does not dictate the pattern of cortical ageing; rather, distinct molecular and network signatures underlie divergent paths of normative decline and compensatory adaptation. Although the present cohort consisted of cognitively unimpaired adults, the morphological resemblance between the normative-ageing subtype and the CDR 1/2 group suggests that coordinated structural change may precede overt cognitive symptoms, bridging normative and pathological ageing (63–65). This interpretation aligns with recent evidence that network-level alterations can emerge before amyloid or tau accumulation in Alzheimer’s disease (66).

In conclusion, our findings support the existence of distinct subtypes of morphometric similarity network organisation associated with specific molecular pathways, revealing network-organisation patterns consistent with normative ageing and compensatory adaptation across the adult lifespan. By integrating morphometric similarity, normative modelling, and gene-expression data, our study provides a biologically informed approach for characterising individual differences in cortical ageing. Spanning early adulthood to late life, this approach identifies distinct network and molecular patterns that may help elucidate mechanisms underlying heterogeneity in healthy brain ageing and their relationship to later-life cognitive impairment.

## Data Availability

All data produced in the present study are available upon reasonable request to the authors

## Author Contributions

Conceptualisation: V.V. (lead), T.R., and A.Y. Methodology: V.V., T.R., A.Y., and V.B. Formal Analysis: V.B. (image processing, SuStaIn, morphometric network analyses), V.V. (genetic analyses). Original Draft Witting: V.V. (lead). Review and Editing: V.V., T.R., A.Y., and V.B. Supervision: V.V. (project lead and overall supervision).

## Data Availability

All data supporting the findings of this study are available from the corresponding author upon reasonable request. The full list of genes, along with derived morphometric similarity network (MSN) measures and subtype classification files, is stored in the [OSF repository folder](https://osf.io/6ra9p/overview) associated with this study.

## Code Availability

The custom MATLAB scripts used for morphometric similarity network analyses and gene–MSN association analyses are available through the project’s [OSF repository folder](https://osf.io/6ra9p/overview). The original SuStaIn implementation is publicly available through [the pySuStaIn repository](https://github.com/ucl-pond/pySuStaIn).

## Additional information

### Supplementary information

The online version contains supplementary material available at *link available upon publication*. Correspondence and requests for datasets should be addressed to Vesna Vuksanovic

## Supplementary Information

### Freesurfer Analysis and Output

All 952 images were preprocessed on the Swansea University High Performance Computer using FreeSurfer (version 6.0) and the *recon-all* command with default parameters for cortical surface reconstruction, ensuring consistency across different software and hardware versions. Additionally, MRI data were corrected for site-specific acquisition differences by including scanning site as a covariate in the analysis, which is a standard procedure followed in multisite studies.

In FreeSurfer, cortical surface area reconstruction is calculated by correcting intensity variations in MRI data, removing non-brain tissue (skull-stripping), segmenting the gray-white matter interface, and separating the hemispheres and subcortical structures. The resulting volume is then filled, giving a representation of the cortical surface. (1). Gray matter volume is calculated by forming an oblique truncated triangular pyramid between matching faces on the white and pial surfaces, which is then divided into three tetrahedra. The total volume the sum of the three tetrahedra (2). The thickness is computed as the average of the thickness between the gray/white boundary surface. The thickness standard deviation is computed based on the variation in these measurements across the vertices of the cortical surface within a specific region of interest (3). Mean cortical curvature is calculated as the average of the principal curvatures at each point on the cortical surface. Gaussian curvature is the product of the two principal curvatures. The folding index (FI) is computed by integrating the product of the maximum principal curvature and the difference between maximum and minimum curvature and dividing by 4p. The Curvature index represents the average amount of curvature per region (4). In short, FreeSurfer outputs were harmonised across datasets following recommendations on image processing and quality control from (5) and taking into account an improved interscanner segmentation stability of the FreeSurfer version 6.0, which was used here (6).

### MRI Datasets

#### The Nathan Kline Institute (NKI) Rock-land Sample

Anatomical T1-weighted MRI data were acquired using standard SIEMENS MAGNETOM Trio-Tim syngo MR B15 scanner using an MPRAGE sequence. The anatomical scan protocol is described in the following summary table ([NKI-PROTOCOL](https://fcon_1000.projects.nitrc.org/indi/pro/nki/NKI_MPRAGE_PROTOCOL.pdf)). Structural images were collected using a three-dimensional high-resolution T1-weighted gradient-echo (MPRAGE) sequence [TR=2.5s, TE=3.5ms, flip angle=8 degrees, matrix size=256×256, voxel size=(1*×*1*×*1mm)^3^, 192 axial oblique slices]. The number of images preprocessed and used from this dataset was N = 119.

**Fig. S1.**
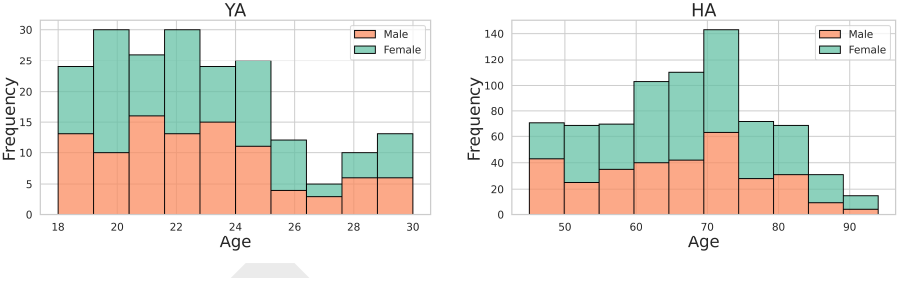
Age histograms of young adults (YA) (n=199) (Left Panel) and healthy ageing(HA) group (n=753) (Right Panel).

#### OASIS

Anatomical T1-weighted MRI data were acquired using a Siemens 1.5T Vision scanner using a Magnetization-Prepared Rapid Gradient Echo (MPRAGE) sequence. Structural images were collected using a three-dimensional high-resolution T1-weighted gradient-echo sequence with the following parameters: (TR): 9.7 ms (TE): 4.0 ms Flip Angle: 10°, Slices: 128 Resolution 256 × 256 (1 × 1 mm) (7). The number of images preprocessed and used from this dataset was N = 357 (OASIS-1) and N = 136 (OASIS-2). (There was 150 images in total in the OASIS-2 dataset, but 14 did not pass QC and were not included.)

#### IEEE

Anatomical T1-weighted MRI data in the Open BHB (Big Healthy Brains) dataset were aggregated from more than 70 acquisition sites across ten publicly available cohorts (e.g., ABIDE-1/2, CoRR, GSP, IXI, Localizer, MPI-Leipzig, NAR, NPC, RBP), encompassing over 5,000 unique healthy control participants ([OPENBHB](https://baobablab.github.io/bhb/dataset)) (8). The number of images preprocessed and used from this dataset was N = 340.

### Genes and MSNs Analysis

Regional gene-expression data were obtained from the Allen Human Brain Atlas (AHBA), which contains microarray measurements from post-mortem healthy adult brains. AHBA expression values were mapped onto the Destrieux Brain Atlas (DBA) regions using anatomical correspondence between the two atlases, following a procedure similar to our previous work (9, 10). Only longevity-associated genes present in both the GenAge and AHBA databases were retained for further analysis. This procedure enabled direct comparison between regional cortical gene-expression patterns and MSN-derived measures at a common anatomical resolution.

**Fig. S2.**
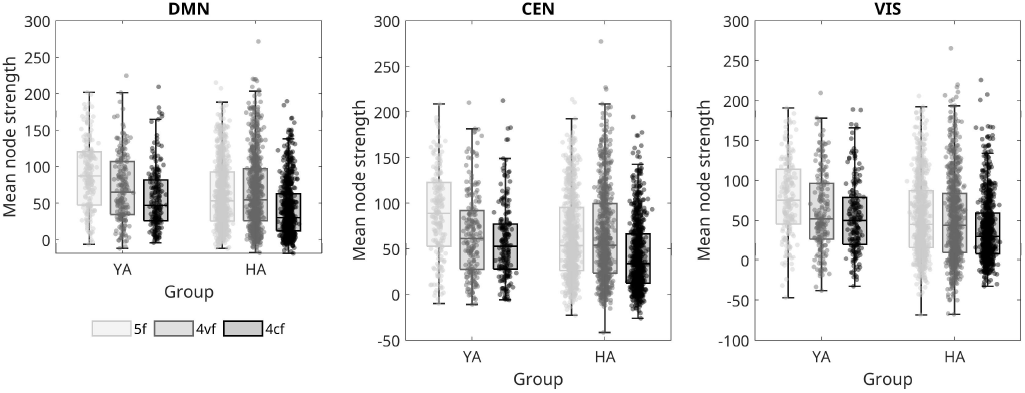
The intra-network node strength for DMN, CEN and VIS networks in the two age groups: healthy young adults and healthy ageing (HA). Statistically significant differences were found between the YA and HA subjects for the 5-feature network in all three networks (DMN:p *<*0.0001; CEN: p *<*0.0001; VIS: p= 0.02), and for the 4-curvature-feature network in the DMN (p= 0.008). The p-values are corrected for multiple comparisons using Bonferroni correction.

**Fig. S3.**
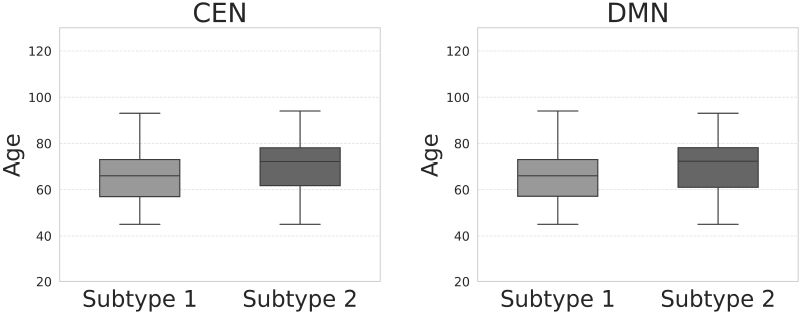
Age boxplots of DMN and CEN of subjects’ subtypes.

To complement the subtype-specific analyses, we additionally examined associations between MSN node strength and longevity-gene expression across all participants pooled together, without stratification into subtypes. This analysis identified 16 genes that remained significant after FDR correction, and their corresponding spatial associations with MSN node strength are shown in Fig. S6. Comparison with the subtype-specific results revealed only four genes (*E2F1, GCLM, GSK3B, MXD1*) that were shared between the pooled analysis and both identified subtypes. Genes shared between pooled and *normative-ageing subtype*: FOXO1, NR3C1, PPARGC1A; and genes shared between pooled and *compensatory subtype*: *DDIT3, FEN1, HSPA8, STUB1, UBB*; and pooled only: *CLU, FOS, GHR, PIK3R1, XRCC*. These findings suggest that while several global gene-MSN associations are present across the entire cohort, the subtype-based analyses reveal additional molecular variation that is not captured when participants are analysed as a single homogeneous group. The pooled analysis recovers additional broad ageing genes (*CLU, GHR, PIK3R1, XRCC, FOS*) that may reflect mechanisms shared across the entire ageing cohort rather than subtype-specific effects.

**Fig. S4.**
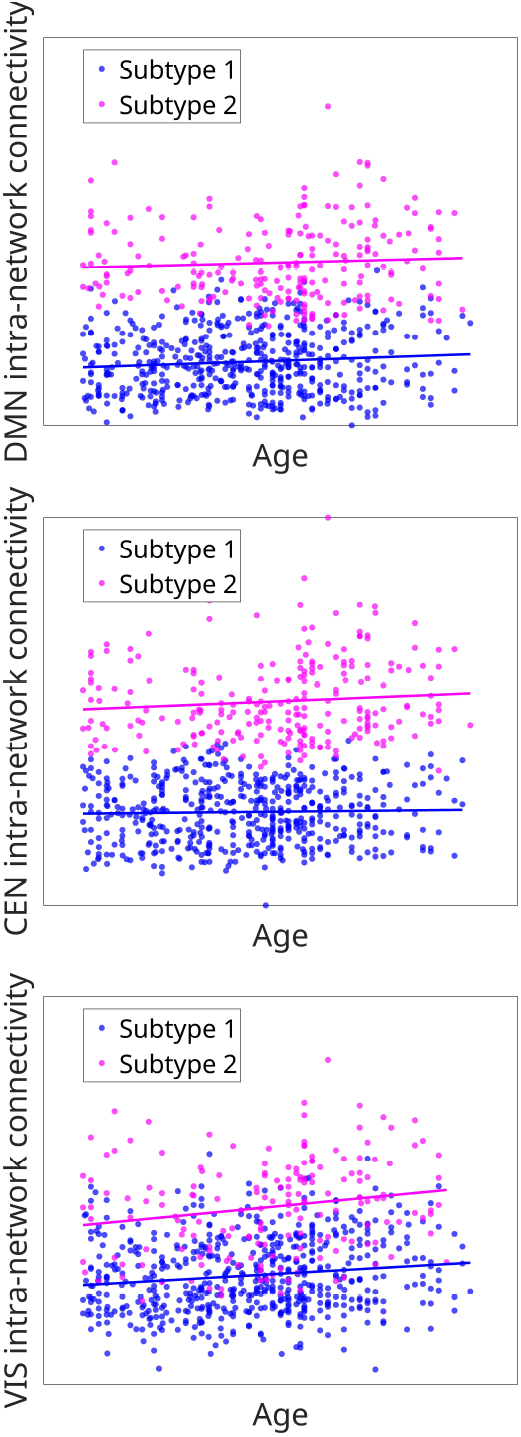
Intra-network connectivity across the default mode (left), central executive (middle), and visual (right) networks as a function of age for two SuStaIn-derived ageing subtypes. Subtype 2 shows consistently higher connectivity than Subtype 1, suggesting a compensatory pattern of network organisation during ageing.

**Fig. S5.**
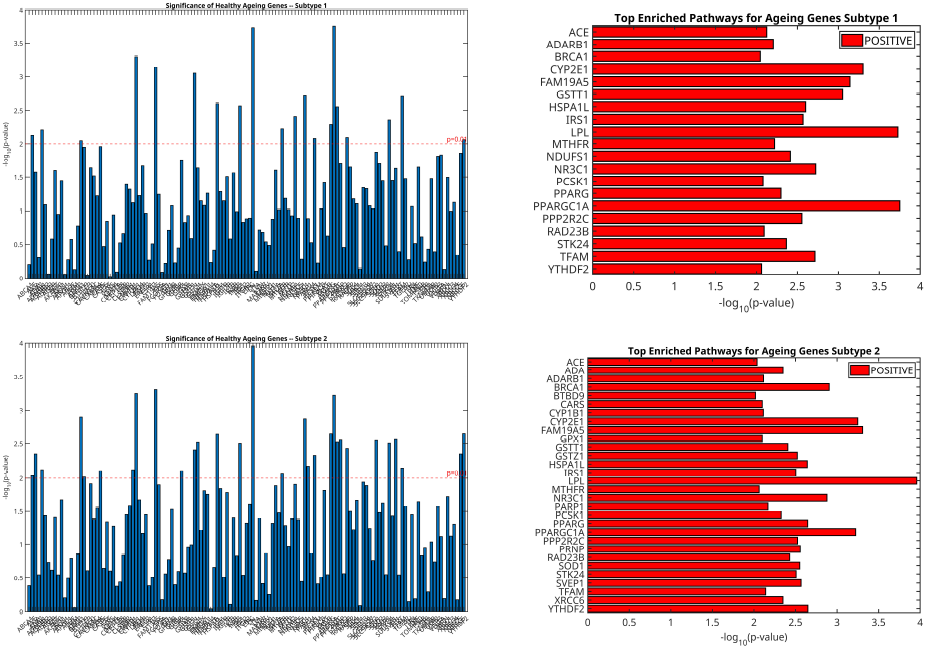
Association between longevity genes and cortical morphometric similarity. (**Top**). Subtype 1 and significant genes (**Right**) Subtype 2 and significant genes. The analysis was performed without corrections for cortical thickness and surface area variations.

**Fig. S6.**
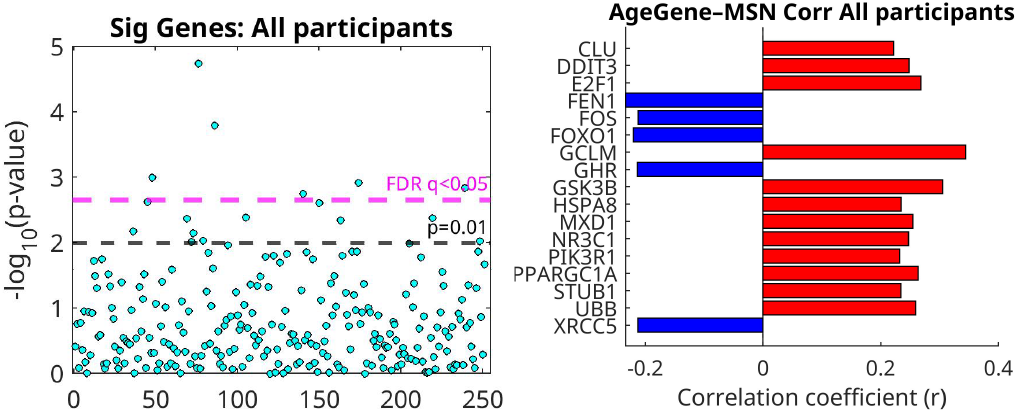
Association between longevity genes and cortical morphometric similarity. Significant genes across all participants. The analysis was performed on all participants (not on each subtype separately) while correcting for cortical thickness and surface area variations.

**Fig. S7.**
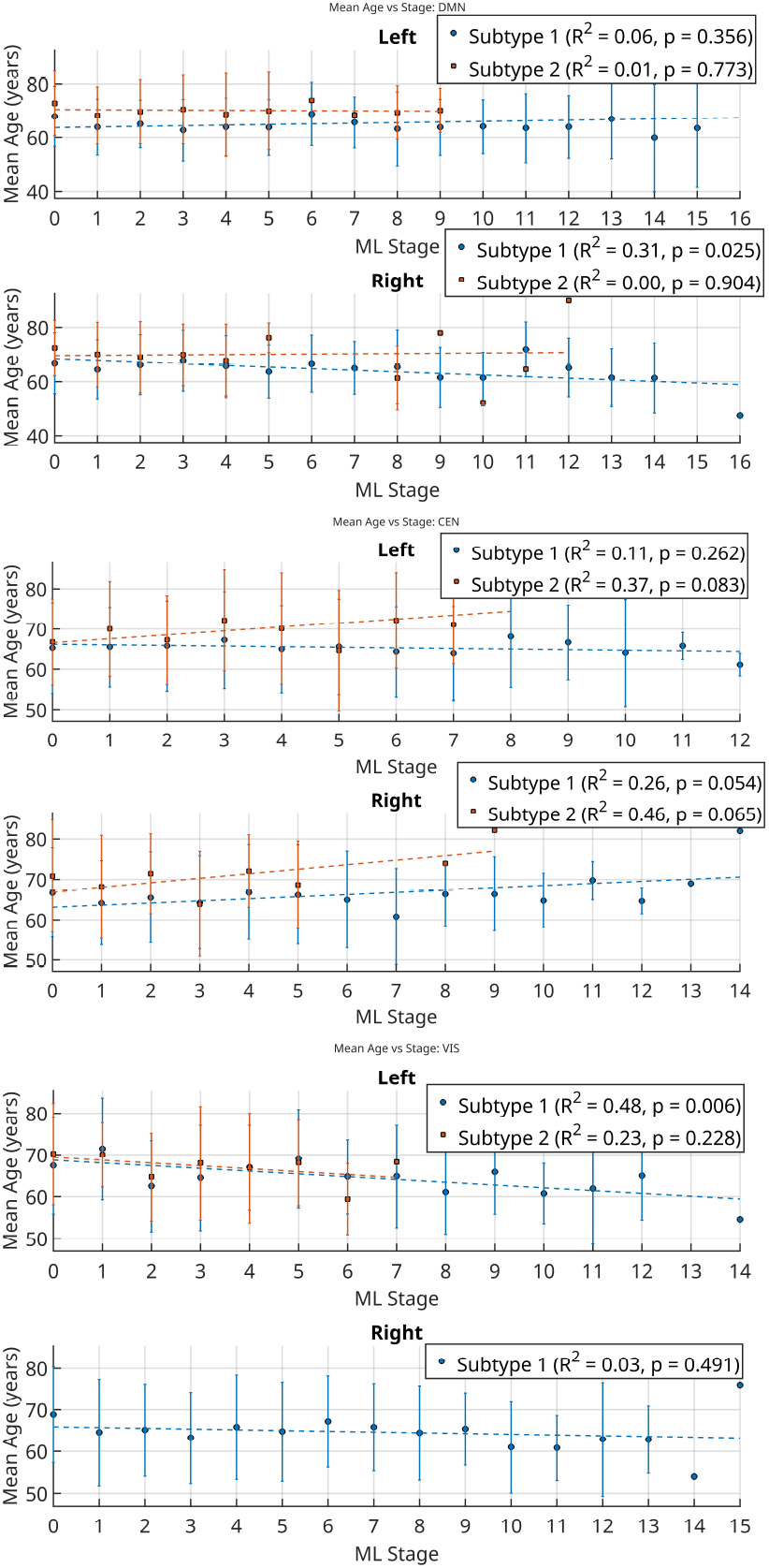
Scatter plots showing mean age across SuStaIn stages for Subtype 1 (normative) and Subtype 2 (compensatory) in the left and right hemispheres of the DMN (top), CEN (middle), and VIS (bottom) networks. Dashed lines represent linear fits, with corresponding *R*^2^ and p-values in the legends. DMN - Default Mode Network; CEN - Central Network; VIS - Visual Network

